# Classification of COVID-19 from Chest X-ray images using Deep Convolutional Neural Networks

**DOI:** 10.1101/2020.05.01.20088211

**Authors:** Sohaib Asif, Yi Wenhui, Hou Jin, Yi Tao, Si Jinhai

## Abstract

The COVID-19 pandemic continues to have a devastating effect on the health and well-being of the global population. A vital step in the combat towards COVID-19 is a successful screening of contaminated patients, with one of the key screening approaches being radiological imaging using chest radiography. This study aimed to automatically detect COVID‐ 19 pneumonia patients using digital chest x‐ray images while maximizing the accuracy in detection using deep convolutional neural networks (DCNN). The dataset consists of 864 COVID‐ 19, 1345 viral pneumonia and 1341 normal chest x‐ray images. In this study, DCNN based model Inception V3 with transfer learning have been proposed for the detection of coronavirus pneumonia infected patients using chest X-ray radiographs and gives a classification accuracy of more than 98% (training accuracy of 97% and validation accuracy of 93%). The results demonstrate that transfer learning proved to be effective, showed robust performance and easily deployable approach for COVID-19 detection.

## I. Introduction

CORONAVIRUS disease (COVID‐ 19) is an infectious disease and looking at the degree of its spread all through the world, it has been declared as a pandemic by the World Health Organization (WHO) on 11th March 2020 [1]. The pandemic pronouncement also stressed the deep worries of the alarming rate of spread and severity of COVID‐ 19. It is the principal recorded pandemic brought about by any coronavirus. It is characterized as a worldwide wellbeing emergency of its time and it has spread everywhere throughout the world. Legislatures of various nations are forcing fringe limitations, flight limitations, social distancing, and expanding consciousness of cleanliness. However, the virus is still spreading at very rapid rate. The majority of the individuals tainted with the COVID‐ 19 experienced gentle to direct respiratory ailment, while some built up destructive pneumonia. There are assumptions that elderly people with basic clinical issues like cardiovascular ailment, diabetes, ceaseless respiratory infection, renal or hepatic maladies and malignant growth are bound to create genuine disease [2]. Until now, there is no particular immunization or treatment for COVID‐ 19. However, there are numerous continuous clinical preliminaries assessing potential medicines. About 5,232,431 infected cases are confirmed in more than 216 countries until 22nd May 2020, where 335,636 deaths, 2,112,080 recovered, 2,739,216 mild and 45,499 critical cases were found [3, 4].

It has been expressed that so as to battle with the spreading of COVID‐ 19 sickness compelling screening of patients and prompt clinical reaction for the contaminated patients is a crying need. The highest quality level screening strategy utilized for testing the COVID‐ 19 patients is the Reverse Transcription Polymerase Chain Response (RT‐PCR) test on respiratory specimens [5]. This procedure is the most generally utilized strategy for testing for COVID‐ 19 identification however is a manual, confused, relentless and time‐consuming process with a positivity rate of only 63% [5]. The other diagnosis tools of COVID‐ 19 can be clinical symptoms investigation, epidemiological history and positive radiographic images (computed tomography (CT)/Chest radiograph (CXR)) as well as positive pathogenic testing. The clinical attributes of serious COVID‐ 19 contamination are that of bronchopneumonia causing fever, hack, dyspnea, and respiratory failure with acute respiratory distress syndrome (ARDS) [6-9]. Promptly accessible and radiological imaging is another major symptomatic instrument for COVID‐ 19. Most of COVID‐ 19 cases have comparable highlights on radiographic pictures including reciprocal, multi‐ focal, ground‐glass opacities with a fringe or back dissemination, primarily in the lower projections, in the early stage and pulmonary consolidation in the late stage [9-15]. Although typical CXR images may help early screening of suspected cases, the pictures of different viral cases of pneumonia are comparative and they overlap with other infectious and inflammatory lung diseases. Therefore, it is hard for radiologists to recognize COVID‐ 19 from other viral pneumonia. The side effects of COVID‐ 19 being like viral pneumonia can at times lead to wrong determination in the present circumstance, where medical clinics are over-burden and working nonstop. Therefore, incorrect diagnosis can prompt a non‐COVID viral Pneumonia being falsely marked as exceptionally suspicious of having COVID‐ 19 and in this manner deferring in treatment with resulting costs, exertion and danger of presentation to positive COVID‐ 19 patients. Currently many biomedical complications (e.g., brain tumor detection, breast cancer detection, etc.) are using Artificial Intelligence (AI) based solutions [16-19]. Deep learning techniques can reveal image features, which are not apparent in the original images. In particular, Convolutional Neural Network (CNN) has been demonstrated amazingly helpful in include extraction and learning and therefore widely adopted by the research community [20]. CNN was utilized to improve picture quality in low‐light pictures from a high‐speed video endoscopy [21] and was additionally applied to distinguish the idea of aspiratory knobs through CT pictures, the conclusion of pediatric pneumonia by means of chest X‐ray pictures, robotized marking of polyps during colonoscopy recordings, cryptoscopic picture acknowledgment extraction from recordings [22-25]. Machine learning techniques on chest X‐Rays are getting popularity as they can be easily utilized with low‐cost imaging techniques and there is an abundance of data available for training different machine‐learning models. Concept of transfer learning in deep learning framework was utilized by Vikash et al. [26] for the recognition of pneumonia utilizing pre‐trained ImageNet models [27] and their ensembles. A customized VGG16 model was used by Xianghong et al. [28] for lung regions identification and various sorts of pneumonia characterization. Wang et al. [29] used a large dataset and Ronneburger et al. [30] used image augmentation along with CNN to show signs of improvement results via preparing on little arrangement of pictures. Rajpurkar et al. [31] announced a 121‐ layer CNN on chest X‐rays to identify 14 distinct pathologies, including pneumonia utilizing an ensemble of different networks. A pre‐trained DenseNet‐ 121 and feature extraction techniques were used in the accurate identification of 14 thoracic diseases in [32]. Sundaram et al. [33] used AlexNet and GoogLeNet with image augmentation to obtain an Area Under the Curve (AUC) of 0.95 in pneumonia discovery.

The test of COVID-19 is currently a difficult task because of inaccessibility of diagnosis system everywhere, which is causing panic. Because of the limited availability of COVID-19 testing kits, we have to depend on different determination measures. Since COVID-19 assaults the epithelial cells that line our respiratory tract, we can utilize X-rays to investigate the strength of a patient’s lungs. The medical practitioner frequently uses X-ray images to analyze pneumonia, lung inflammation, abscesses, and enlarged lymph nodes. And almost in all hospitals have X-ray imaging machines, it could be possible to use X-ray’s to test for COVID-19 without the dedicated test kits. Again, a drawback is that X-ray examination requires a radiology master and takes huge time, which is valuable when people are sick around the world. Therefore, developing an automated analysis system is essential to save medical professionals valuable time.

Recently, several groups have described deep‐learning based COVID‐ 19 pneumonia detection techniques [34-36]. Shuai et al. [35] used deep learning techniques on CT images to screen COVID‐ 19 patients with an accuracy, specificity and sensitivity of 89.5%, 88% and 87% respectively. Linda et al. [34] presented a DCNN, called COVID‐Net for the detection of COVID‐ 19 cases from the chest X‐ray images with an accuracy of 83.5%. Ayrton [36] used a small dataset of 339 images for training and testing using ResNet50 based deep transfer learning technique and reported the validation accuracy of 96.2%. In this study, we have developed an automatic detection of COVID-19 using a DCNN based Inception V3 model and Chest X-ray images. This paper proposes advanced deep learning approach to predict the COVID-19. The proposed work is implemented with TensorFlow and Inception V3 pre-trained models that was trained to classify normal, viral and COVID‐ 19 pneumonia images and tested on Chest X-ray images and obtained more than 98% of classification accuracy.

### A. Problem Definition

In order to control the spread of COVID-19, a large number of suspected cases need to be screened for proper isolation and treatment. Pathogenic research facility testing is the indicative best quality level however it is tedious with noteworthy bogus negative outcomes. Quick and precise analytic strategies are desperately expected to battle the sickness. In light of COVID- 19 radiographical changes in X-ray pictures, we meant to build a deep learning method that could extract COVID-19’s graphical features so as to give a clinical analysis in front of the pathogenic test, thus saving critical time for disease control. In this paper, (DCNN) [37], a machine learning classification technique is used to classify the Chest X-ray images. As accuracy is the most significant factor in this issue, by taking a more prominent number of pictures for training the network and by increasing the number of iterations, the DCNN accuracy can be improved. Tensor Flow is a large-scale machine learning system developed by Google [38] and Inception V3 is Google’s CNN architecture [39]. Here, the DCNN algorithm is executed with Tensor Flow and Inception V3.

## II. MATERIALS AND METHODS

DCNN typically perform better with a larger dataset than a smaller one. Transfer learning can be beneficial in those applications of CNN where the dataset is not large. The idea of transfer learning uses the trained model from large datasets such as ImageNet [40] is used for application with comparatively smaller dataset. This eliminates the requirement of having large dataset and also reduces the long training period as is required by the deep learning algorithm when developed from scratch [40].

### A. Collection of Dataset

In this study, 315 chest X-ray images of COVID-19 patients have been obtained from the open source GitHub repository shared by Dr. Joseph Cohen [41]. This repository is containing chest X-ray/CT images of mainly patients with acute respiratory distress syndrome (ARDS), COVID-19, Middle East respiratory syndrome (MERS), pneumonia, severe acute respiratory syndrome (SARS). In addition, 330 COVID‐ 19 positive radiographic images (CXR and CT) were carefully chosen from Italian Society of Medical and Interventional Radiology (SIRM) COVID‐ 19 DATABASE [42]. Out of 330 radiographic images, 70 images are chest x‐ray images and 250 images are lung CT images. This database is updated in a random manner and until 29 March 2020, there were 63 confirmed COVID‐ 19 cases were reported in this database. In addition, 2905 chest X-ray images were selected from COVID- 19 Radiography Database [43]. Out of 2905 radiographic images, there are 219 COVID-19 positive images, 1341 normal images and 1345 viral pneumonia images.

### B. Image Pre-processing

One of the significant phases in the data preprocessing was to resize the X‐Ray images as the image input for algorithm were different. We implemented some image pre-processing technique to increase the performance to our system by speeding up training time. First, we resized all our images to 299×299×3 to increase processing time and also to suitable in Inception V3. In the image preprocessing step, we need to label the data since the learning technique of convolution neural network fits into administered learning in machine learning.

### C. Image augmentation

CNN needs a sufficient amount of data to achieve excellent performance. We apply data augmentation techniques to increase the insufficient data in training, and the techniques used include vertical flip, horizontal flip, noise, translation, blur and rotate the image 60 °, 90 °, 180 °, 270 °. Therefore, the initial dataset consisting of 864 COVID-19 images, 1341 normal images, and 1345 viral pneumonia images was expanded to a total of 8,640 COVID-19 images, 13,410 normal chest X-ray images, and 13,450 viral pneumonia images.

**TABLE I.**
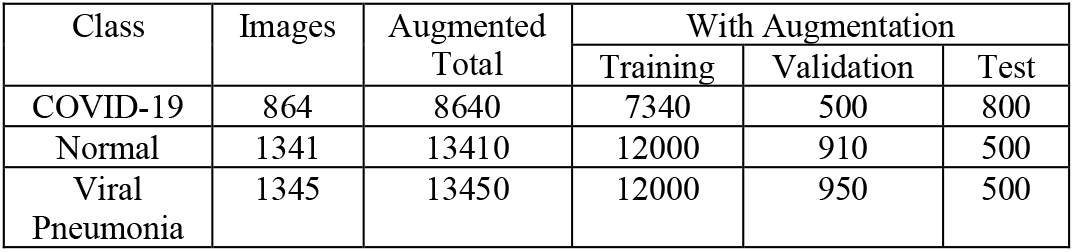
Details of Training, Validation and Test set.

### D. Transfer Learning

Transfer learning is a machine learning technique [40] which is based on the concept of reusability Transfer learning is often used with CNN in the way that all layers are kept except the last one, which is trained for the specific problem. This technique can be particularly useful for medical applications since it does not require as much training data, which can be hard to get in medical situations. In the analysis of medical data, one of the biggest difficulties faced by researchers is the limited number of available datasets. Deep learning models often need a lot of data. Labeling this data by experts is both costly and time consuming. The biggest advantage of using transfer learning method is that it allows the training of data with fewer datasets and requires less calculation costs. With the transfer learning method, which is widely used in the field of deep learning, the information gained by the pre-trained model on a large dataset is transferred to the model to be trained.

Fig. 3 defines the Inception V3 model which performs convolution, pooling, softmax and fully connected procedures. Here a pre-trained neural network established for one task can be utilized as the initial point of another task. The Inception-v3 architecture comprises two fragments:

**Fig. 1.**
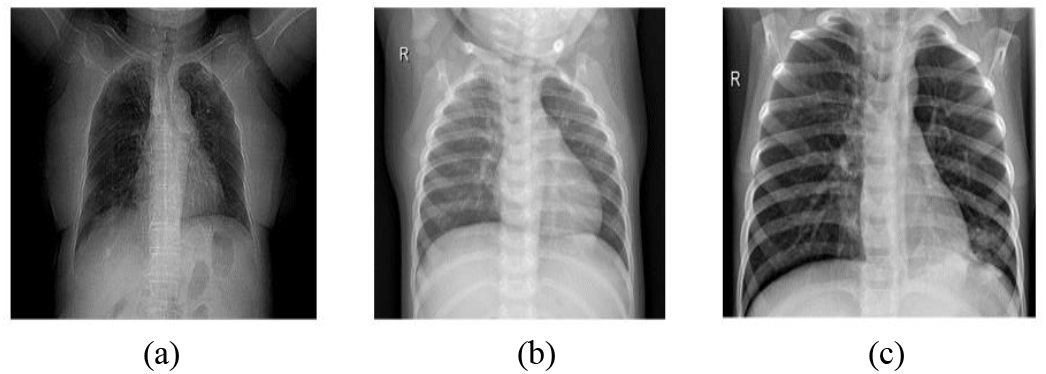
Example chest radiography images of: (a) COVID-19 Viral Infection (b) Non COVID-19 infection (c) Viral Pneumonia.

**Fig. 2.**
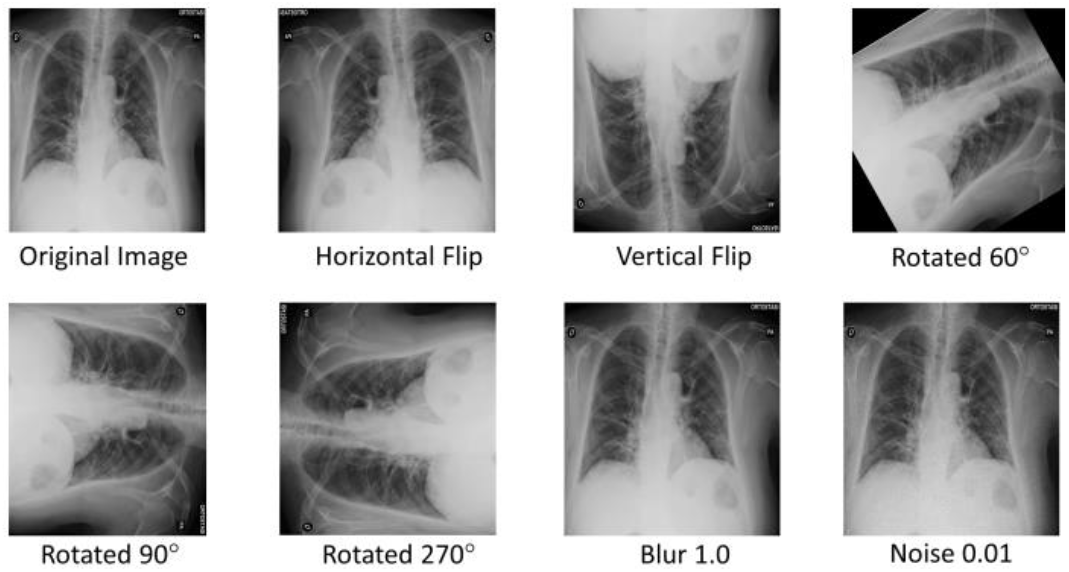
Example of augmented images by rotating, flipping, blur and noise.

**Fig. 3.**
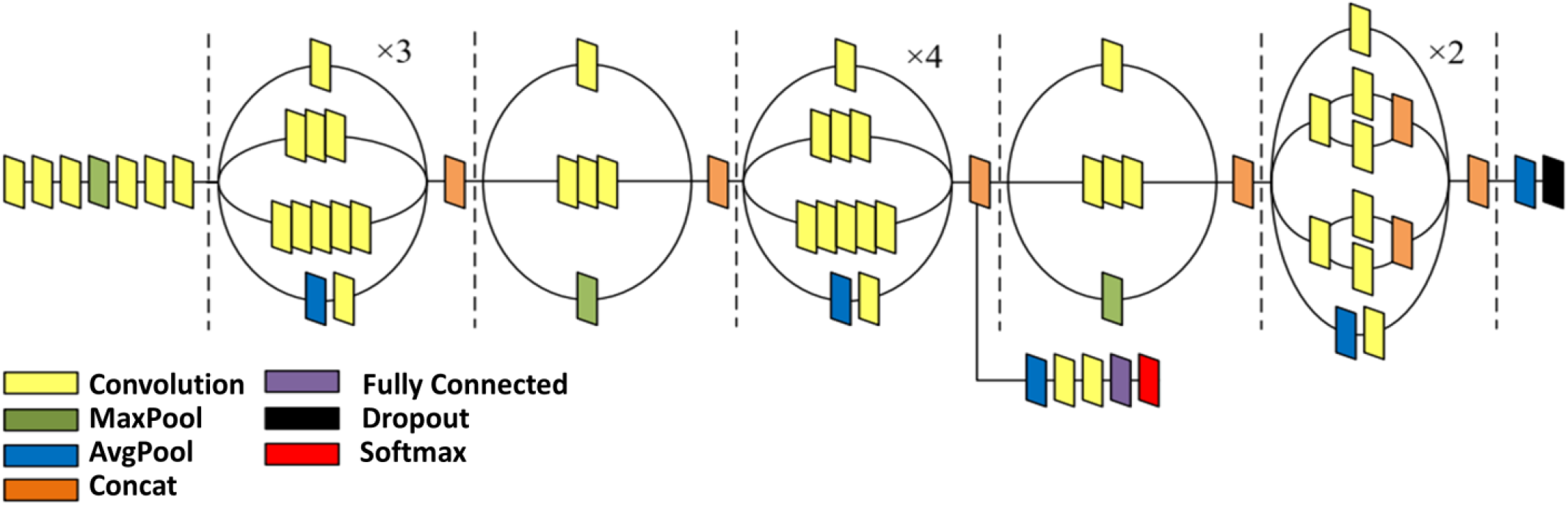
Inception V3 Architecture.

I. Use the feature extraction section of the convolutional neural network.
II. Classification section utilizing fully-connected and softmax layers.

### E. TensorFlow

It is an artificial neural network there are more than three layers, shown in Fig. 4. It has single input, single output and many invisible layers [38]. To use transfer learning for classifying chest X-ray images, we used the TensorFlow library [38] to load the Inception V3 model on our local machine, retrain it on the chest X‐ray dataset and then classify new images to be one of the three categories normal, viral pneumonia and COVID‐ 19. It is a deep learning framework established by Google that can control all neurons (nodes) in the system and has a library appropriate for image processing. Neural network weights can be changed to improve performance.

**Fig. 4.**
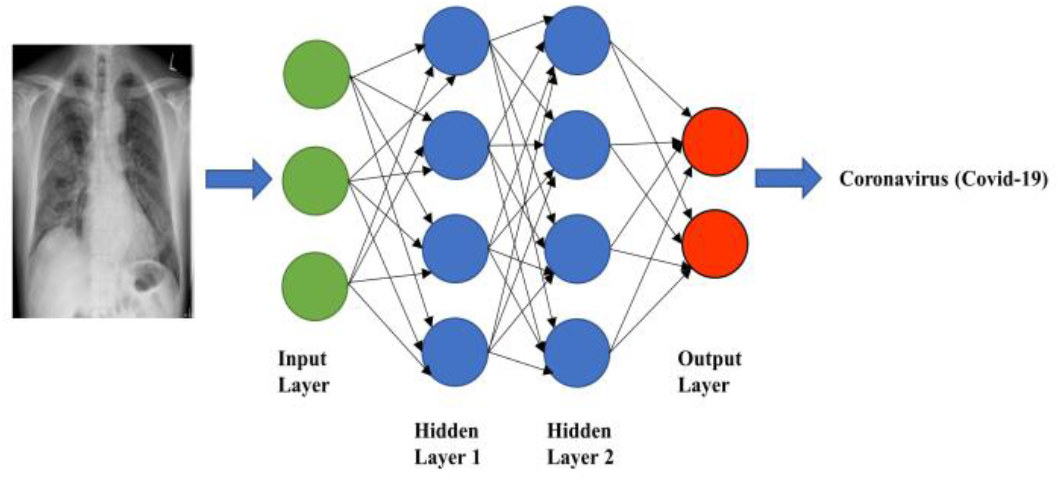
Neural network classifier.

### F. Proposed Architecture

Fig. 5 provides a step-by-step procedure for the proposed work model. The steps for the projected classification architecture are as follows:

I. Recursively perform convolution and pooling on images.
II. Apply drop out and fully connected. Now the image must be classified according to the labelled training class.

Convolution is a gradual process. Extracts various features of input. Each kernel is responsible for producing output function. Low-level features of the image, such as edges, lines and corners are determined by the lower layer, and the higher-level features are extracted by the higher layer.

Pooling is applied to make the features obtained from convolution robust against noise. Pooling layers are usually of two types namely, average pooling and max pooling. It is basically a dimensionality reduction or feature extraction step. A simple example of max and average pooling is shown in Fig. 6.

In this study, we built DCNN based InceptionV3 model for the classification of COVID-19 Chest X-ray images to normal, viral pneumonia and COVID-19 classes. In addition, we applied transfer learning technique that was realized by using ImageNet data to overcome the insufficient data and training time. The schematic representation of conventional CNN including InceptionV3 model for the prediction of COVID-19 patients, viral pneumonia and normal were depicted in Fig. 7. Chest X-ray images are taken as input, Inception V3 is applied, convolution, pooling, softmax, and fully connected processes are performed. Upon completing these tasks, they are classified according to different training modules and eventually classified as normal, viral pneumonia and COVID-19 classes. Inception V3 is one of the states of art architectures in image classification challenge. The best network for medical image analysis seems to be the Inception V3 architecture and it preforms better than even the more recent architectures. So, we selected Inception V3 model that is implemented using TensorFlow and hence the retraining is done with TensorFlow. The steps for classification using the proposed work are follows:

**Fig. 5.**
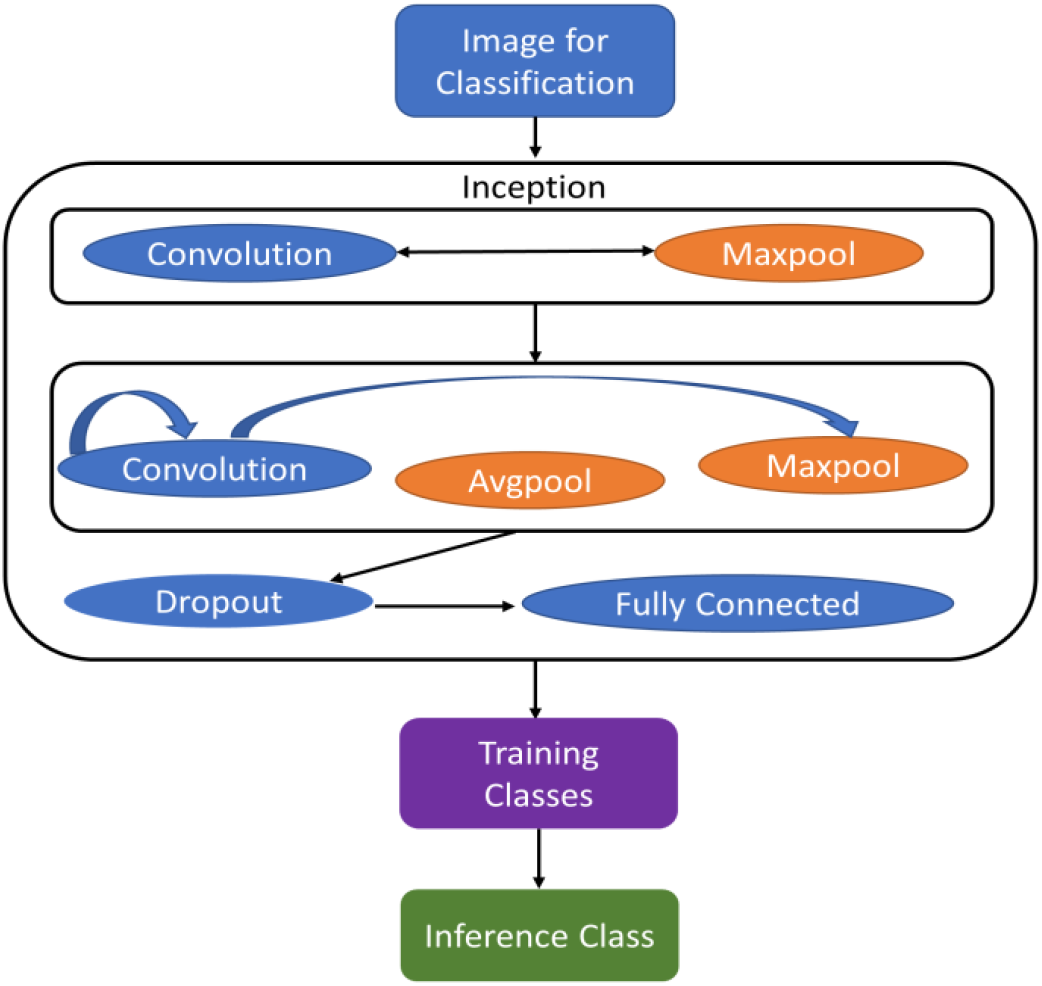
Proposed work architecture.

**Fig. 6.**
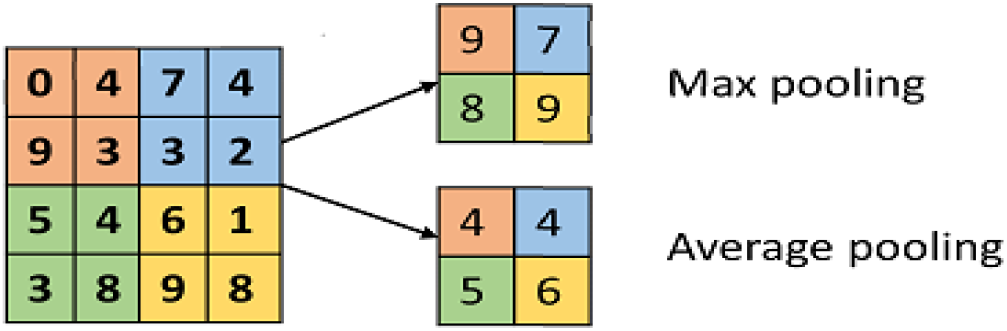
Example of Max and Average Pooling.

**Fig. 7.**
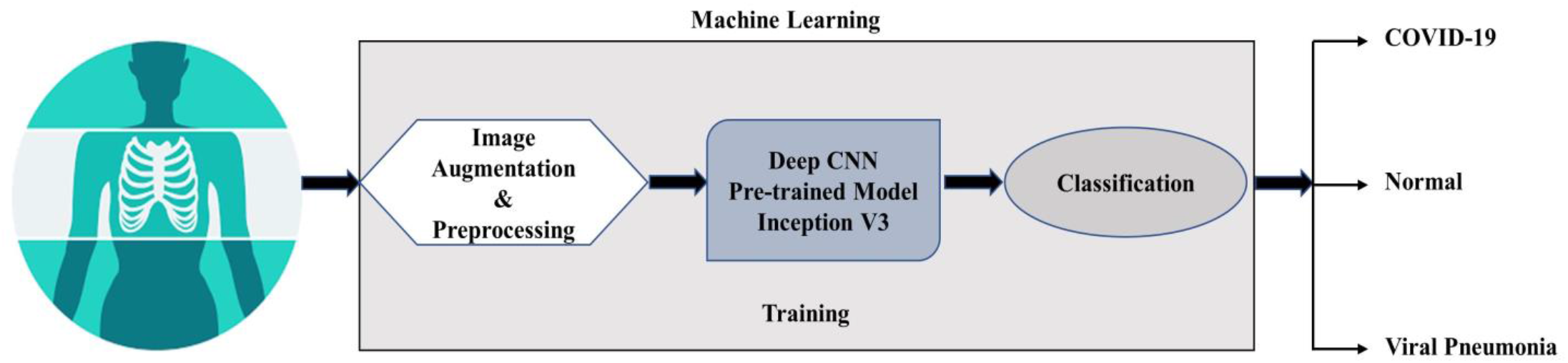
Schematic representation of pre-trained model for the prediction of COVID-19 patients, normal and viral pneumonia.

Algorithm TensorFlow Classification Step 1: Start

Step 2: Create list of images // start training the model

Step 3: Provide a directory for storing the bottleneck value of each image

Step 4: Provide inference to the images // to create bottleneck values

Step 5: Create a folder for all images of bottleneck values

Step 6: Generate bottleneck values for each individual image

Step 7: Create new softmax layers and fully connected layers // end of training

Step 8: Test new image // input chest x-ray image to get the result

Step 9: Finish

## III. Results and Discussion

In this study, we had presented a novel method that could screen COVID-19 fully automatically by DCNN. Chest X-ray images have been used for the prediction of COVID-19 infected patients. Popular pre-trained model Inception V3 has been trained and after training, the model was experienced on chest X-ray images of COVID-19, normal and viral pneumonia that are not used in the training phase. We have obtained the best performance as a classification accuracy of more than 98%. The experimental work is done by connecting the docker to the virtual box. The neural network is trained to create bottleneck values. As shown in Fig. 8. (a), create a bottleneck for individual image and save it to a folder. After all the bottlenecks have been created, the last few layers of the model are trained. We have set the training steps to 4000, which is sufficient to generate sufficient outcomes. There are a series of accuracy, cross-entropy, and verification accuracy as shown in Fig. 8. (b). Training accuracy indicates a correctly labeled share of the current training image. Cross-entropy indicates how finely the training is performed on the cross-entropy loss function, and validation accuracy is related to training accuracy. Training accuracy increased as training proceeds after 4000 iterations, the final test accuracy was 96%.

**Fig. 8.**
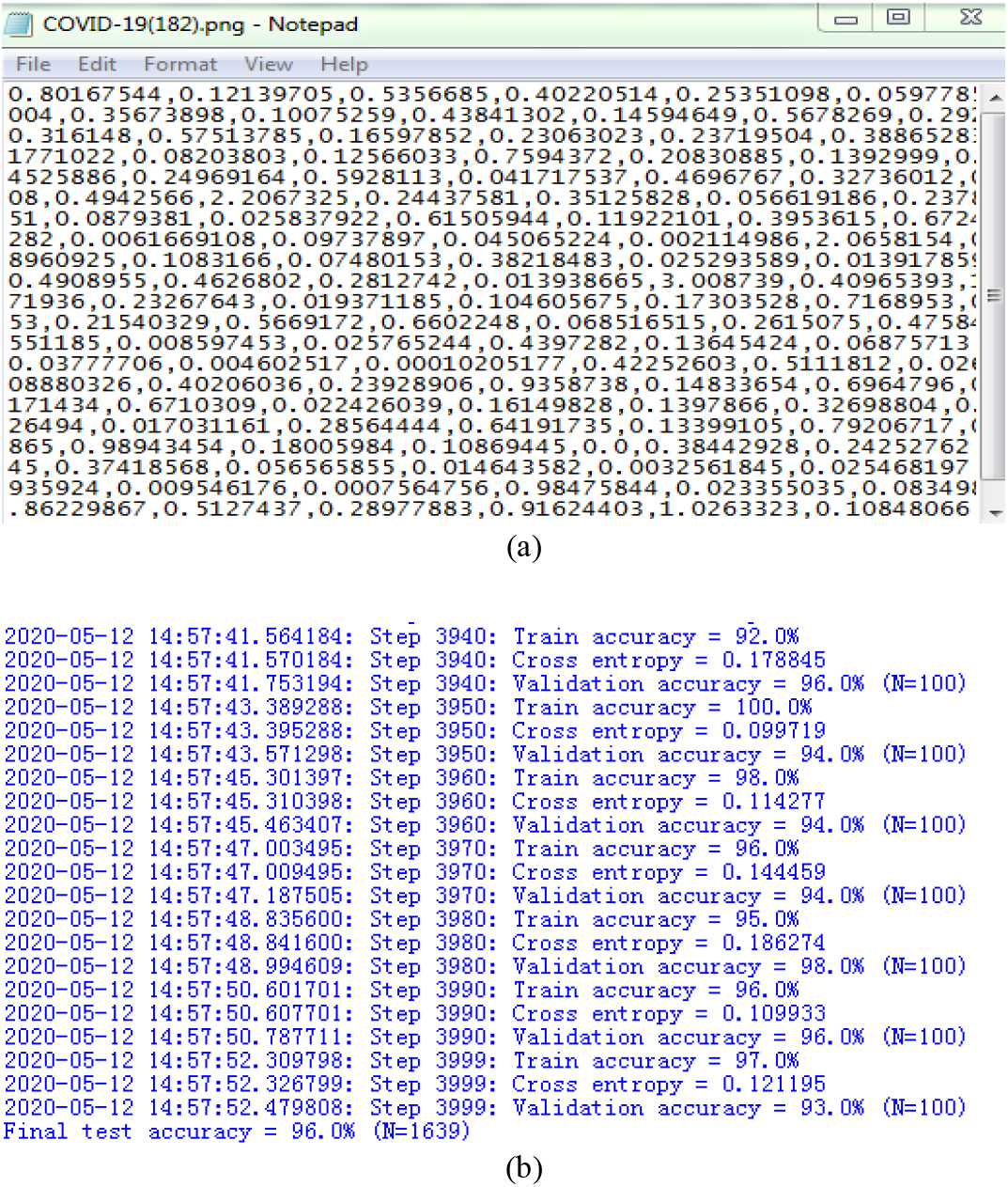
(a) Generation of Bottleneck values (b) Final test accuracy of the retraining.

### A. Training accuracy & Cross-entropy

We measured the training accuracy and cross-entropy during the training steps as shown in Fig. 9.

**Fig. 9.**
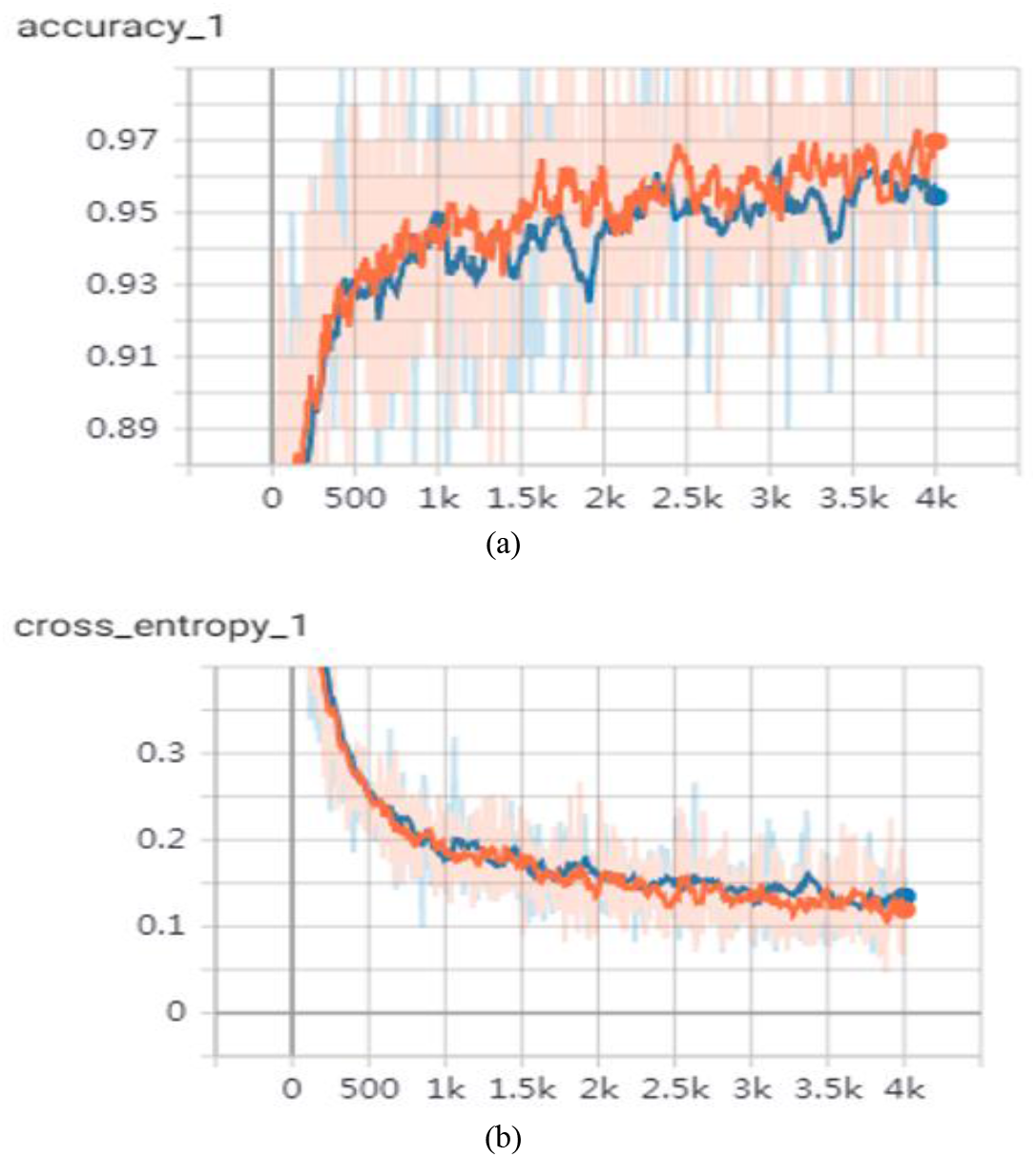
(a) Training and validation accuracy of model and (b) cross entropy. Orange curve indicates the training cross-entropy/accuracy and blue curve indicates the validation cross-entropy/accuracy.

The training accuracy Fig. 9. (a) illustrates the percentage of the images used in the current dataset that were labelled with the correct class and the validation accuracy Fig.9. (a) shows the percentage of randomly selected correctly labeled images from a different set. The core difference is that the accuracy of training is based on the images that the network can learn, so the network can over adapt to the noise in the data. Cross entropy Fig. 9. (b) is a loss function which gives a sight into how well the process of learning is progressing, lower numbers are better here. To provide more human-interpretable explanations, we conducted several experiments on the chest X-ray images to evaluate the classification performance of the network investigated, let’s consider the following examples.

Example 1: the CXR image is classified to contain a confirmed COVID-19 case with a probability of 99.59%, the true class is COVID-19, as shown in Fig. 10. (a).

**Fig. 10.**
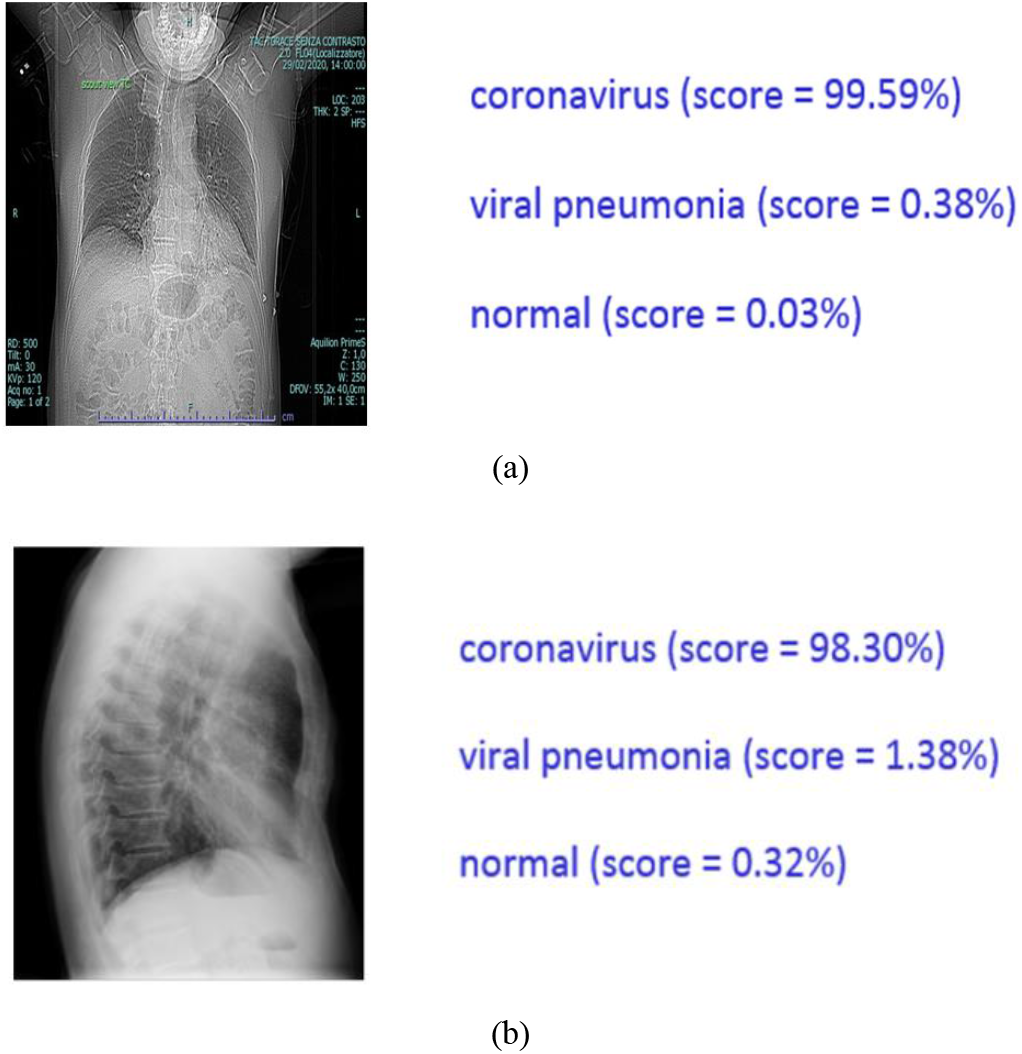
Example CXR images of COVID-19 cases from 2 different patients (A) Classification performance obtained by testing our trained architecture with first CXR image (B) Classification performance obtained by testing our trained architecture.

Example 2: the CXR image is classified to contain a confirmed COVID-19 case with a probability of 98.30%, the true class is COVID-19, as shown in Fig. 10. (b).

Fig. 10. shows the results when the sample test image was taken. Since COVID-19 has a higher score compared to normal and viral pneumonia, the test image is classified as COVID-19. It can be concluded that the proposed technique can classify COVID-19 X-ray images very reliably.

## Data Availability

Datasets used in the experiments were obtained from J. C. Monteral. (2020). COVID Chest X-ray Database (https://github.com/ieee8023/covid-chestxray-dataset), Italian Society of Medical and Interventional Radiology (https://www.sirm.org/category/senza-categoria/covid-19/) and COVID-19 Radiography Database (https://www.kaggle.com/tawsifurrahman/covid19-radiography-database). All data needed to evaluate the conclusions in the paper are present in the paper. Additional data related to this paper may be requested from the authors.

https://www.sirm.org/category/senza-categoria/covid-19/

https://www.kaggle.com/tawsifurrahman/covid19-radiography-database

## IV. Conclusion

Early prediction of COVID-19 patients is important to avoid spreading the disease to different people. In this study, we proposed a deep transfer learning-based approach the use of chest X-ray images obtained from COVID-19 patients, normal and viral pneumonia for automatic detection of COVID-19 pneumonia. The proposed classification model for the detection of COVID-19 achieved more than 98% accuracy. In the light of our findings, it’s far believed that it’s going to help medical doctors to make decisions in scientific practice due to the high overall performance. In order to come across COVID-19 at an early stage, this study gives insight on how deep transfer learning methods can be used. COVID‐ 19 has already become a danger to the world’s healthcare system and thousands of people have already died. Deaths were initiated by way of respiration failure, which ends up in the failure of other organs. Since a big range of sufferers attending out‐ door or emergency, doctor’s time is limited and computer-aided-analysis can save lives via early screening and proper‐ care. Inception V3 model exhibits an excellent performance in classifying COVID‐ 19 pneumonia by effectively training itself from a comparatively lower collection of images. We believe that this computer-aided diagnostic tool can significantly improve the speed and accuracy of diagnosing cases with COVID‐ 19. This could be highly useful in a pandemic, where the burden of disease and the need for preventive measures do not match the availability of resources.

## Notes

### Competing Interest Statement

The authors have declared no competing interest.

### Funding Statement

This work was supported by the National Key R&D program of China (Grant No. 2019YFA0706400, 2019YFA0706402), the Pre-research Funds for Equipment of China (no. 61409220115), the Natural Science Foundation of China (61275179, 60970815), the Natural Science Basic Research Plan in Shaanxi Province of China (program no. 2016JM8047), the Fundamental Funds for the Central Universities (no. xjj2015112), ShaanXi Province administration of traditional Chinese medicine (15-ZY036), and Xi'an medical university state fund cultivation project (2016GJF01).

